# Identification of implications of m6A regulators and autophagy-associated genes for prognosis in ovarian cancer

**DOI:** 10.64898/2026.06.25.26356535

**Authors:** Yanru Chen, Xiaoni Yu, Weixin Chu, Shipeng Shang, Ningning He, Li Guo

**Author notes:** Correspondence (N.H.); (L.G.).

## Abstract

The most prevalent RNA alteration in the mammalian genome is N-6-methylenediosine (m6A). There is mounting evidence linking dysregulation of m6A regulatory factors and alterations in m6A levels to the development, course, or prognosis of ovarian cancer. Genes having prognostic value were screened using the univariate, multifactorial, and Least Absolute Shrinkage Selection Operator (LASSO) Cox regression analyses. Important genes’ m6A expression in clinical material was verified by real-time fluorescent quantitative polymerase chain reaction (RT-qPCR). In present study, all 23 regulators were significantly differentially expressed in ovarian cancer tissues. LASSO regression analysis screened for 10 key genes associated with both autophagy and m6A. A risk score was constructed and nomogram was developed to forecast the prognosis of ovarian cancer patients. Additionally, individuals with ovarian cancer were classified as high-risk or low-risk; and the low-risk group might be more likely to benefit from immunotherapy. RT-qPCR was used for the bioinformatics study of human ovarian cancer and normal tissues. Lastly, PLK2 and LEPR were confirmed to be associated with tumorigenesis in scRNA-seq. The risk score established by m6A and autophagy can be used to predict prognosis and susceptibility to anticancer drugs in patients with ovarian cancer.

## 1. Introduction

One of the most common gynecological cancers worldwide is ovarian cancer. In 2020, there were 314,000 reported new instances and 207,000 deaths from ovarian cancer(1). Registry data from the US and UK reveal that approximately 1 in 6 women succumb within 90 days post-diagnosis, highlighting the high morbidity and mortality associated with late-stage presentation. These outcomes underscore the need for timely and appropriate treatment, which could significantly reduce the disease’s impact and improve patient survival(2). Due to the absence of reliable early screening indicators, late-stage detection accounts for the majority of ovarian cancer patients’ dismal survival rates(3). While surgery and chemotherapy are standard treatments, drug resistance frequently leads to fatal outcomes(4). A diverse collection of malignant tumors affecting the ovary, fallopian tube and peritoneum make up ovarian cancer(5). The primary cause of the variation in ovarian cancer patients’ prognoses is tumor heterogeneity.

In eukaryotic messenger RNAs, the nitrogen atom of the sixth adenine (A) undergoes a common methylation alteration known as N6-methyladenosine (m6A). In the 3’ untranslated regions (3’-UTRs) of the mRNA and close to stop codons, this alteration is frequently observed (6). Its occurrence is regulated by specific proteins known as m6A regulators. The m6A regulators were divided into three categories, including writers (ZC3H13, METTL16, METTL14, METTL3, VIRMA, RBM15B, RBM15 and WTAP), readers (FMR1, YTHDF3, YTHDF2, YTHDF1, YTHDC2, YTHDC1, RBMX, HNRNPC, HNRNPA2B1, LRPPRC, IGFBP3, IGFBP2 and IGFBP1) and erasers (FTO and ALKBH5). There is mounting evidence linking alterations in m6A levels and m6A regulator dysregulation to the onset, progression, or prognosis of ovarian cancer(7). METTL3 influenced m6A modification of TRPC3 to mediate ovarian cancer metastasis(8). Ovarian cancer cells’ stemness was diminished by FTO via lowering the m6A levels of PDE1C and PDE4B (9). Abnormal expression of m6A regulators has been observed in various cancers, suggesting a potential role in tumor development(10) In ovarian cancer, ALKBH5 is over-expression, and its silencing has been shown to reduce EGFR/PI3K/AKT signaling activity. This downregulation leads to the suppression of ovarian cancer cell proliferation and invasion(11, 12). Programmed cell death pathways is a process of cell suicide, which is necessary to maintain organisms homeostasis(13). Multiple programmed cell death including necroptosis, pyroptosis, autophagy and ferroptosis had special role on health and disease(14). Pyroptosis was facilitated citric acid by CASP4/TXNIP-NLRP3-Gesdermin-d pathway, which may be helpful in the therapy of ovarian cancer(15). In this study, an association between m6A and autophagy was found in ovarian cancer. A risk score was developed to evaluate the prognosis of patients with ovarian cancer, taking into account both autophagy and m6A. The risk score has a part in anticipating the response to immunotherapy and the drug sensitivity of chemotherapy, and can guide ovarian cancer patients’ personalized medicine.

## 2. Materials and Methods

### 2.1. Data collection and preprocessing

The GEO and GTEx databases, as well as the Xena public data centers, provided the RNA-seq data and copy number variation (CNV) profiles for both normal and cancerous ovarian tissues. 379 samples of ovarian cancer from the TCGA database and 88 samples of normal ovarian tissue from the GTEx database were gathered in total. Furthermore, 130 ovarian cancer samples from the GEO dataset (GSE138866) were included. The m6A methylation profiling of ovarian cancer tissues were download from GSE119168.

### 2.2. Expression and mutation analysis of m6A regulators

A p<0.05 criterion was applied to the Wilcoxon rank sum test in order to ascertain the difference in gene expression between ovarian cancer and ovarian tissue. Mutation analysis of m6A regulators was carried out using Rpackage “maftools”.

### 2.3. Gene set enrichment analysis

Programmed cell death-associated genes were collected from Zou’s research 39376916 Programmed cell death score was calculated by “GSVA” R package. The association between the programmed cell death score and the m6A regulators was examined using Pearson correlation analysis. A correlation was considered significant if the correlation coefficient (|r|) exceeded 0.3 and the p-value fell below 0.05.

### 2.4. Generation of the risk score

Firstly, autophagy-associated genes and m6A regulators were merged to screen for ovarian cancer prognosis-related genes. A 2:1 ratio of the TCGA dataset was randomly assigned to the training and testing sets. Key genes were identified in training sets using Least Absolute Shrinkage and Selection Operator (LASSO) regression analysis. After selecting ten genes and their coefficients, and the risk score was calculated using the following formula:

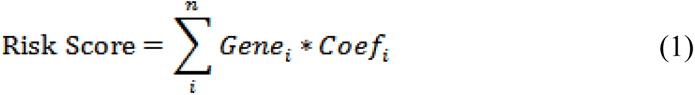

where each selected gene’s expression level is denoted by 〖Gene 〗_i, and the matching LASSO coefficient is indicated by〖Coef〗_i.

### 2.5. Drug sensitivity and immunotherapy Analysis

Drug sensitivity was estimated by using the R package “oncoPredict”. The drug sensitivity indicator known as 50% of cellular growth inhibition (IC50) was utilized. To calculate TMB of ovarian cancer samples, synonymous variant and intron variant was excluded. Finally, the number of mutations on the million-base was calculated as the TMB of the sample. Additionally, the TIDE score of ovarian cancer samples was assessed using the Tumor Immune Dysfunction and Exclusion (TIDE) database.

### 2.6. Single cell data processing

scRNA-seq of ovarian cancer samples was downloaded from GEO (GSE184362) and processed with the Seurat R package (v4.3.0). For quality assurance, cells with less than 200 expressed genes were filtered out, as well as genes present in less than three cells and those with a mitochondrial gene percentage more than 20% were eliminated. Principal component analysis (PCA) was performed on the 2000 most variable genes, using the first 30 principal components for UMAP, which was segmented with a resolution of 0.6. Based on the SingleR package and expression level of marker, cell identities were annotated.

### 2.7. Real-time fluorescent quantitative polymerase chain reaction (RT-qPCR)

Archived fallopian ovarian tissue samples from ovarian cancer patients were retrieved from the Qingdao Municipal Hospital tissue bank for this study. The samples were accessed for research purposes on 3 March 2024. All samples were fully de-identified prior to analysis, and the authors had no access to any information that could identify individual participants during or after data collection. The use of these archived samples was approved by the Institutional Review Board of Qingdao University (approval no. QDU-HEC-2024261), which waived the need for additional informed consent. Total RNA was isolated from the fallopian tubes and ovaries of ovarian cancer patients using Trizol reagent (Vazyme, Nanjing, China) and the fallopian tubes were used as controls. Using the RNA DNA HiScript II qRT SuperMix (Vazyme, Nanjing, China), the extracted RNA was reverse-transcribed into cDNA in accordance with the manufacturer’s instructions to guarantee product quality. A Roche LightCycler 960 real-time PCR machine and ChamQ SYBR qPCR Master Mix (Vazyme) were used to perform the qPCR studies. An internal control for gene quantification was established using GAPDH. During the RT-PCR analysis, biological and technical duplicates of every gene were run at least three times. Table 1 lists the primer sequences that were used.

**Table 1.**
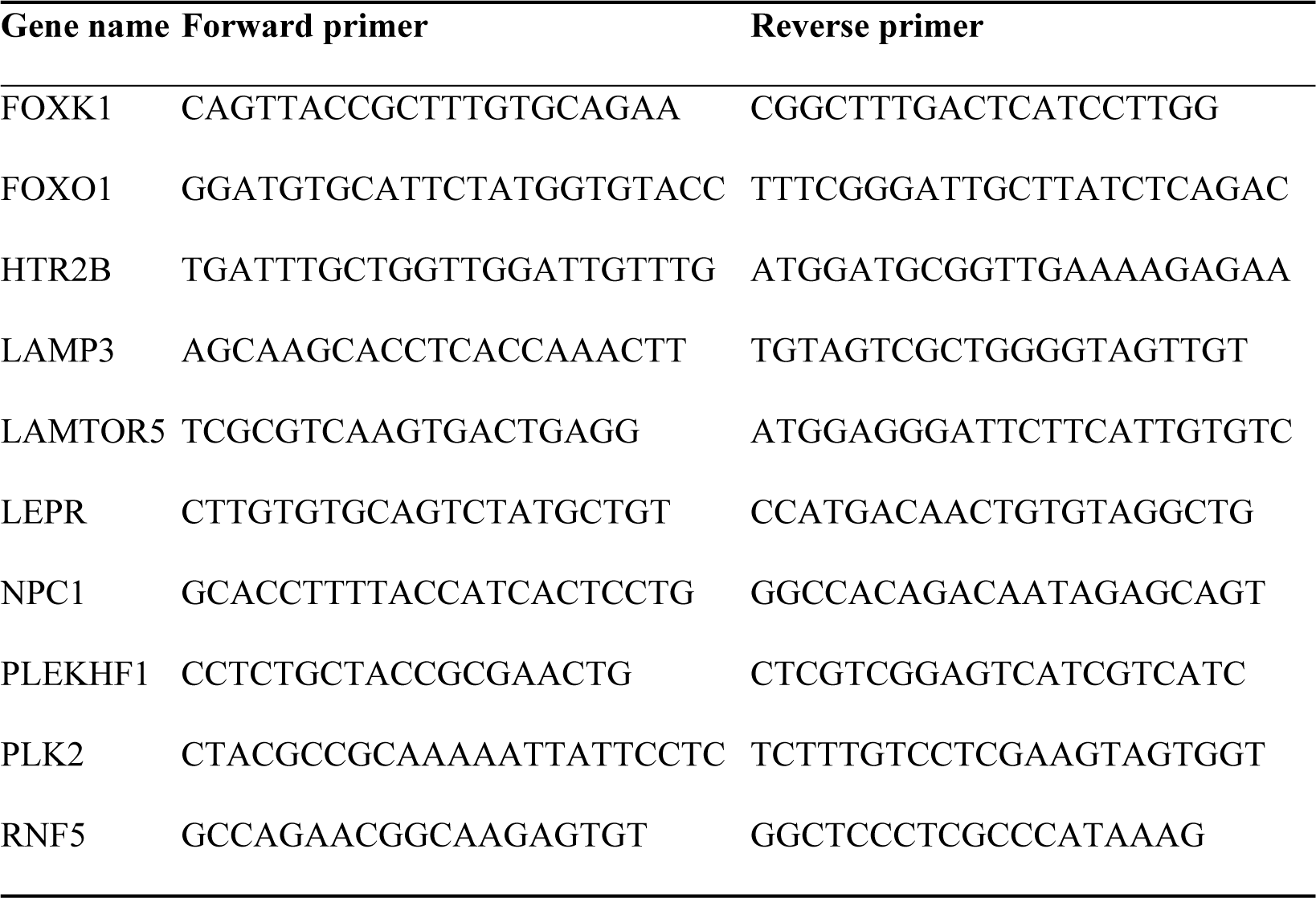
Primer sequences.

### 2.8. Statistical analysis

Correlation analysis was carried out using pearson method. To evaluate the differences between the two groups, the Wilcox rank sum test was utilized. R software (v4.2.2) was used for all statistical analysis, and significantity was defined as p < 0.05.

## 3. Results

### 3.1. Multi-omics characteristics of m6A regulators in ovarian cancer

The m6A regulates gene expression and controls several other critical biological processes in malignant tumor and its level is regulated by m6A regulators. To explore regulatory patterns of m6A regulators, expression level of 23 m6A regulators was compared between ovarian cancer and normal tissues. YTHDF1, RBM15B, YTHDF2, YTHDF3, RBM15, IGFBP2, ALKBH5, LRPPRC and ZC3H13 were up-regulated in ovarian cancer. FMR1, IGFBP3, METTL14, VIRMA, RBMX, WTAP, METTL16, HNRNPC, FTO, YTHDC1, METTL3, YTHDC2, HNRNPA2B1 and IGFBP1 were down-regulated in ovarian cancer (Figure 1A). In ovarian cancer, the overall mutation rate of 23 m6A regulators is comparatively modest (Figure S1). Subsequently, we calculated the CNV frequencies for these m6A-related regulators in ovarian cancer samples, aiming to identify genes that exhibit significant amplification or deletion events. Among m6A regulatory genes, YTHDF1, YTHDF3, VIRMA, METTL3 and HNRNPC exhibited higher rates of amplification while YTHDF2 and WTAP showed a relatively high frequency of deletion (Figure 1B). Further, we analyzed the influence between m6A regulators in ovarian cancer patients and found most m6A regulators showed positive correlation in the expression level. YTHDF3 and VIRMA had the most significant correlation with a correlation coefficient of 0.7. In addition, the influence of 23 factors on the prognosis of cancer patients was evaluated. WTAP and VIRMA were found be risk factors for ovarian cancer patients by using survival analysis (Figure 1C).

### 3.2 Correlation analysis between m6A regulators and autophagy

m6A mediates downstream targets to govern programmed cell death and it is regulated by regulatory factors, such as writers and erasers, and recognition factors, such as readers(16). We collected 12 programmed cell death gene sets for GSVA to study their association with m6A regulators in ovarian cancer. After calculating the correlation between the scores of 12 programmed cell death and the m6A regulators, and it was found that there was a strong positive link between the expression levels of m6A genes and programmed cell death. Among them, autophagy was most obviously associated with m6A regulators, which 18 out of 23 m6A regulators showed significant positive correlation with autophagy (Figure 2A). To further verify the association between autophagy and m6A regulators, m6A profiling of ovarian cancer tissues was collected from GEO database(17). 1814 genes were differential expression between ovarian tissues and normal tissue and 51 of them were autophagy-associated genes (Figure 2B). Specifically, 14 genes were significant up-regulated in m6A level and 37 genes were significant down-regulated (Figure 2C).

### 3.3 Construction of risk score in ovarian cancer

Autophagy and m6A were associated with prognosis of cancer patients(18, 19). Thus, expression level of autophagy-associated genes and m6A regulators was used to evaluate prognosis of ovarian cancer patients. 36 genes were associated with ovarian cancer prognosis by using univariate cox analysis. 32 genes were risk factor of overall survival for ovarian cancer and 4 genes were protective factor (Figure S2A). Ovarian cancer samples were split into training set and testing set at random according to the ratio of 2:1. LASSO model was used to select key genes and verify performance of genes by using ten-fold cross-validation (Figure S2B-C). To build the risk score, 10 key genes were chosen using a Lasso regression model combined with ten-fold cross-validation on the training set (Figure 3A). The risk score was calculated using the following formula: risk score = 0.0003* FOXK1 + 0.1224* FOXO1 + 0.0640* HTR2B - 0.1469* LAMP3 - 0.1422* LAMTOR5 + 0.0917* LEPR + 0.0248 * NPC1 + 0.1400 * PLEKHF1 + 0.0074 * PLK2 - 0.0435* RNF5. In train set and test set, the groups were categorized into high-risk and low-risk according to the median risk score (cutoff of train set: −0.3654487, cutoff of test set: −0.3733432) and had a significant difference in survival (Figure 3B-C). We computed the risk score for ovarian cancer patients from GSE138866 dataset to evaluate its robustness. The survival outcomes between high-risk and low-risk groups showed a significant difference (Figure 3D). Additionally, the key genes exhibited differential expression at the protein level between ovarian cancer and normal tissues (Figure S3).

Further, we evaluated predictive performance of risk score’s ability in terms of ovarian cancer patients’ 1-, 3-, and 5-year survival. The 1-year, 3-year, and 5-year survival times for ovarian cancer were predicted by the risk score with AUC values of 0.5998, 0.5956, and 0.7005, respectively (Figure 4A). Age, stage, and risk score were combined to construct a nomogram that was used to predict the survival of ovarian cancer patients (Figure 4B). The calibration curve demonstrated that, overall, the nomogram’s anticipated survival was basically consistent with the actual survival (Figure 4C-E).

**Figure 1.**
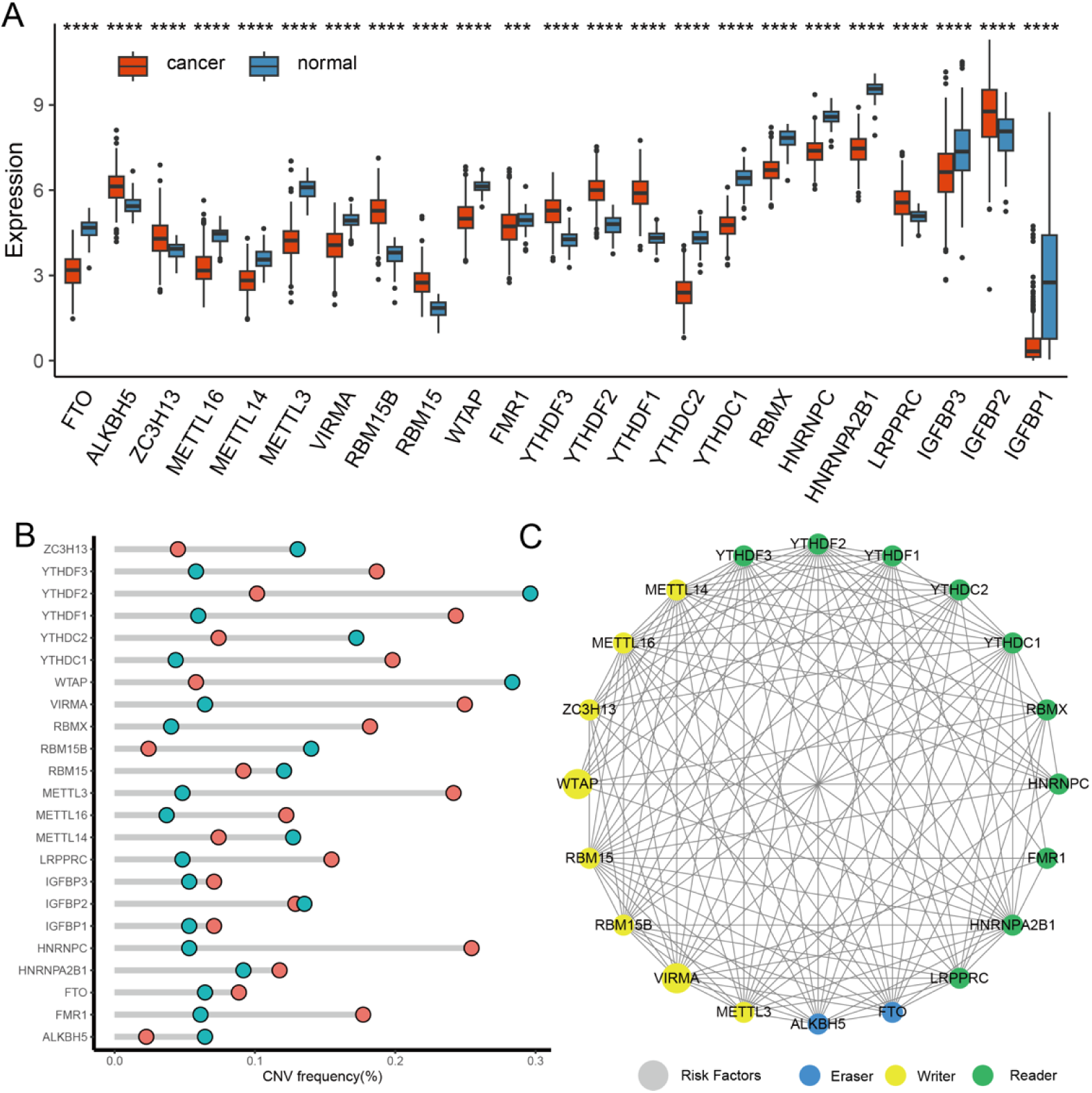
Genetic alterations in m6A-associated genes. (A) Genetic variations in m6A-associated genes. (B) CNV frequency in genes linked to m6A. Turquoise represents amplification, orange represents deletion. (C) PPI network: interactions between m6A-related regulators. Circle size indicates the survival impact of each gene, with larger circles signifying a more significant effect; the circle colors differentiate gene functions: green for recognition proteins (readers), yellow for methyltransferases (writers), and blue for demethylases (erasers); the lines connecting the genes show genetic interactions.

**Figure 2.**
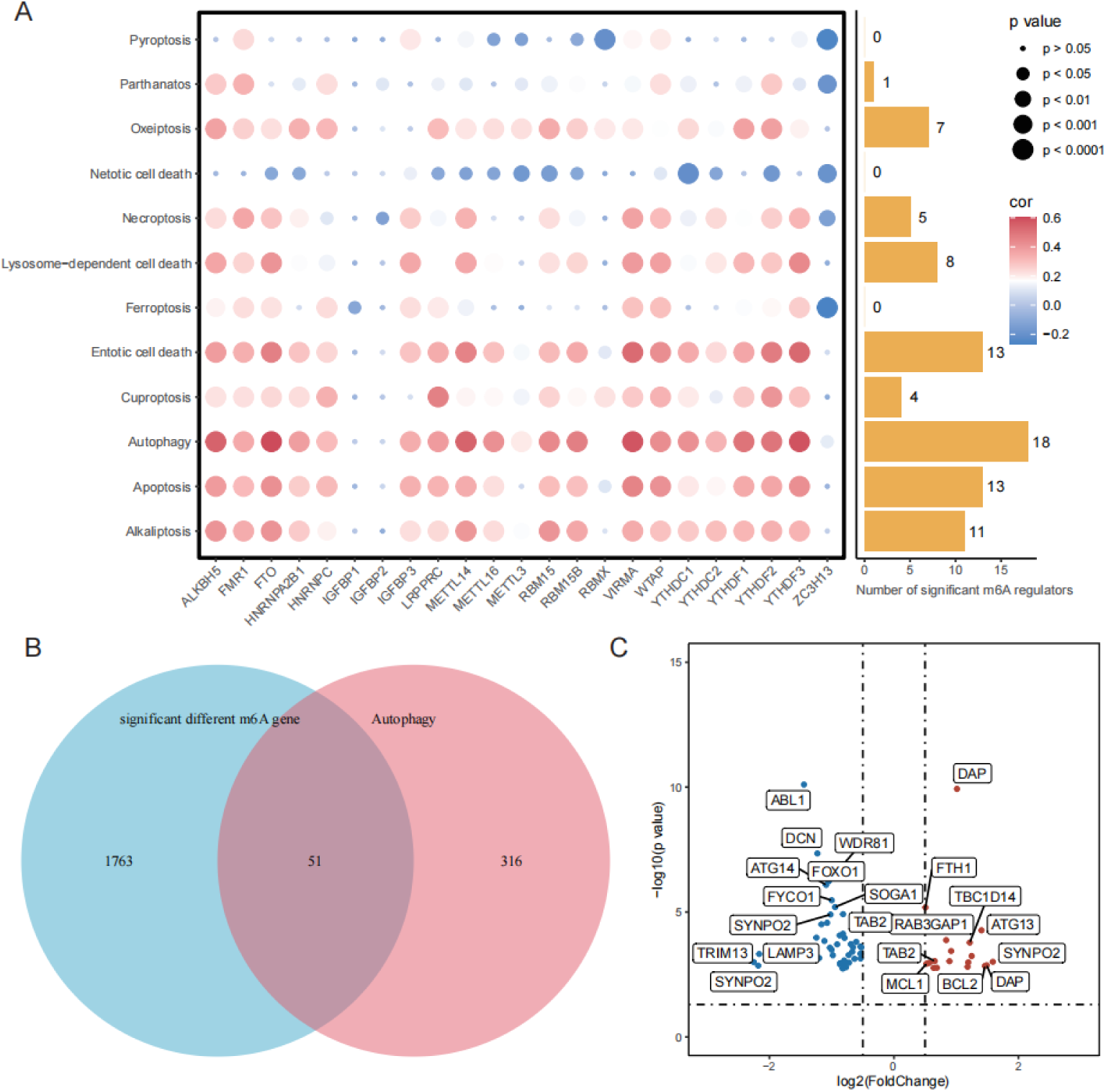
The relationship between autophagy and m6A regulators in ovarian cancer: (A) GSVA enrichment analysis showed m6A regulators were positively enriched in the autophagy. (B) The Venn diagram shows the overlapping genes generated by the intersection of significant different m6A genes and autophagy gene set. (C) Volcano plot showed that 51 differential genes associated with autophagy exhibited varying expression levels in m6A level.

**Figure 3.**
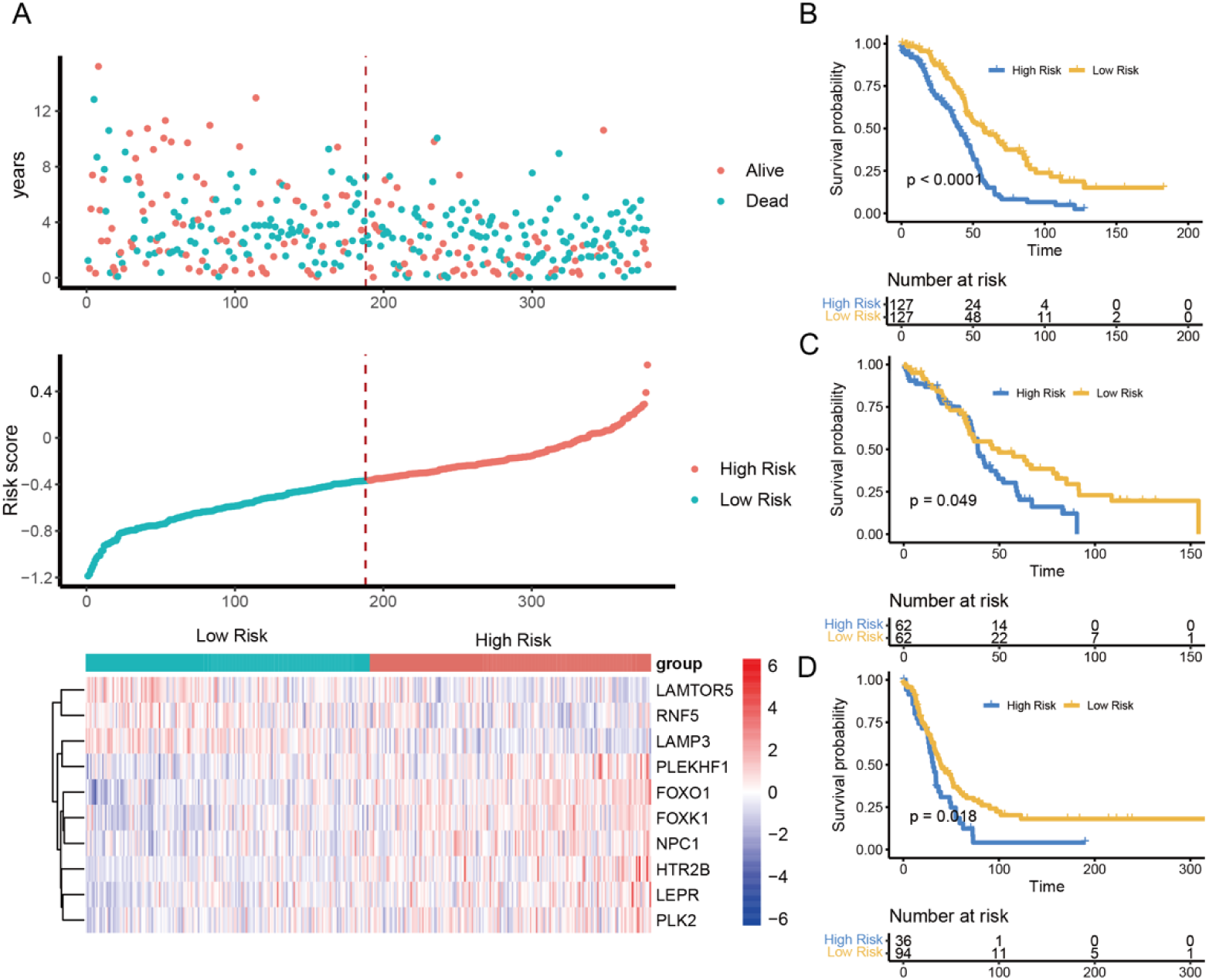
(A) The risk score, along with patient survival status distribution among low-and high−risk groups, and heat maps showing the expression of 10 m6A-related genes. (B-D) Kaplan–Meier survival analysis for patients with high or low risk scores in the train set (B), test set (C) and GSE138866 cohort (D).

**Figure 4.**
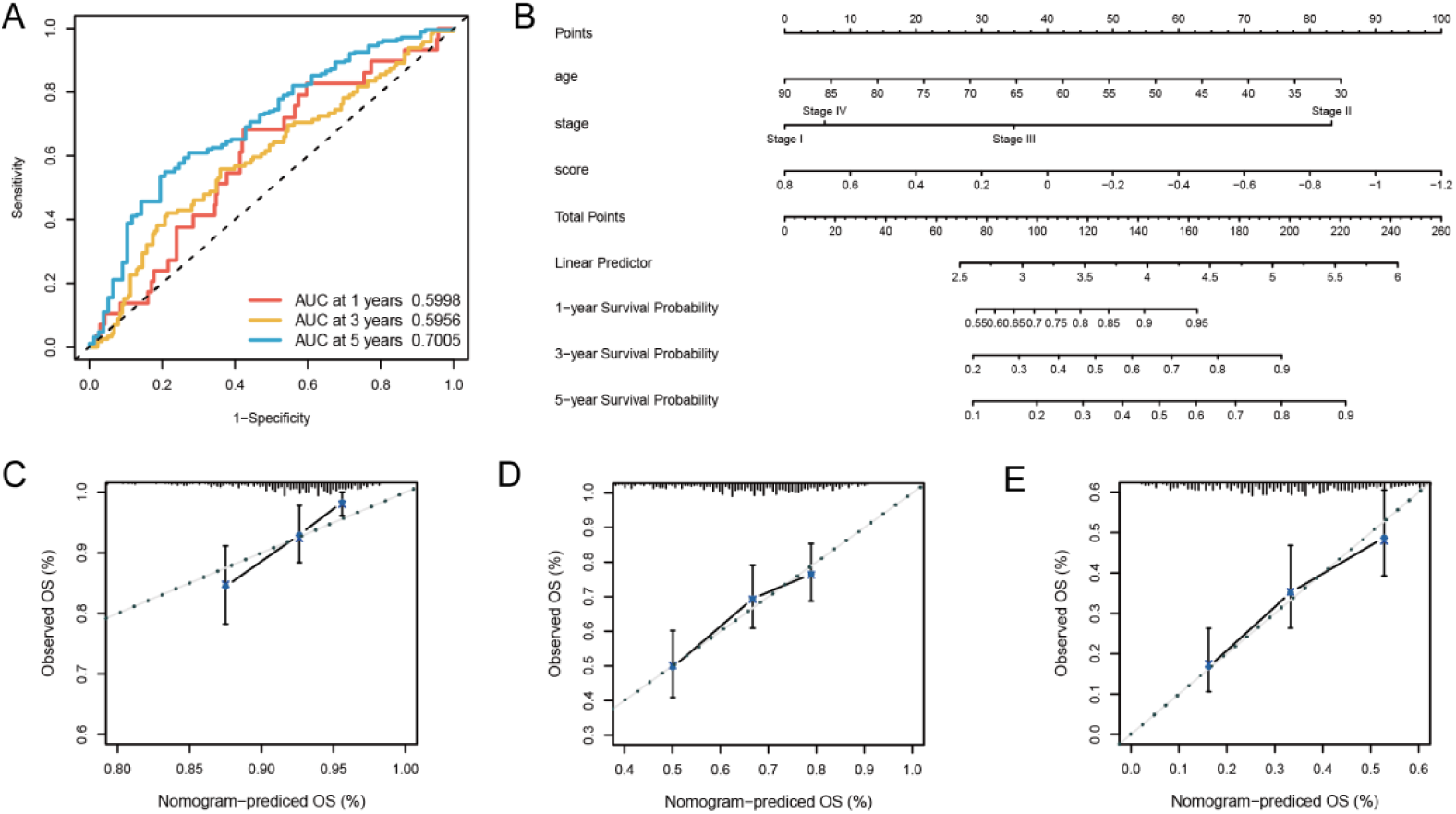
Nomogram analysis. (A) ROC curve was used to evaluate the prognostic risk signature’s prediction ability. (B) A nomogram was constructed with stage, age, and risk score to estimate the likelihood of ovarian cancer at 1, 3, and 5 years. (C-E) The calibration curves for the nomogram for 1-year (C), 3-year (D), and 5-year (E) ovarian cancer.

### 3.4 High-risk and low-risk groups’ therapeutic responses

We calculated anti-cancer drug sensitivity of multiple drugs for ovarian cancer in order to investigate how risk factors affect the clinical treatment outcomes. Docetaxel and Paclitaxel were commonly used drugs of ovarian cancer and high risk group had higher drug sensitivity of Docetaxel and Paclitaxel than low risk group (Figure 5A-B), which meant these chemotherapeutic medications may benefit the high-risk population better. In addition, HAVCR2, LAYN, LAG3 and VDR exhibited differential expression between high and low-risk groups (Figure 5C). Both the TIDE score and the tumor mutation load were high in the low-risk group (Figure 5D-E). The above results indicated that low-risk group may be more likely to benefit from immunotherapy.

### 3.5 Validation of the key feature genes by RT-qPCR

To further validate the expression of key feature genes, tumor tissue and paratumor tissue from ovarian cancer patients were collected. PLEKHF1, FOXO1, LEPR, HTR2B, NPC1, PLK2, FOXK1 and LAMP3 were chosen for the study and their expression was confirmed using RT-qPCR (Table 1). Results showed that LAMP3 was expressed at significantly higher levels in adjacent normal tissues compared to ovarian tumor tissues, whereas PLEKHF1, FOXO1, LEPR, HTR2B, NPC1, PLK2, and FOXK1 exhibited increased expression in ovarian tumor tissues compared to normal tissues (Figure 6).

### 3.6 scRNA-seq analysis reveals expression of key genes in ovarian cancer

After quality control, 88,068 cells from the scRNA-seq data of 6 ovarian cancer samples and 6 paratumor samples identified 21 clusters by the UMAP. According to the annotation of the marker genes, we classified 21 cell clusters into eight cell types, comprising T cells, B cells, NK cells, Endothelial cells, Epithelial cells, Monocyte, Neutrophils and Frbroblasts (Figure 7A-B). Epithelial cells are mainly derived from ovarian cancer tissue (Figure 7C). HTR2B, LAMP3, LAMTOR5, LEPR, NPC1, PLK2 and RNF5 has high expression level in Cancer tissue samples compared to paratumor samples (Figure 7D). PLK2 and LEPR was significant high expressed in epithelial cells compared to other cell type. Further, PLK2 positive/negative group and LEPR positive/negative group were divided according to whether the cells expressed PLK2 and LEPR. Functional enrichment analysis showed that PLK2 was associated with epithelial mesenchymal transformation, apoptosis and p53 pathway. LEPR was associated with fatty acid metabolism and hypoxia (Figure 7E-H).

## 4. Discussion

Ovarian cancer has a very high incidence in female. Ovarian cancer is often referred to as a "silent killer" because it is frequently diagnosed at advanced stages due to its typically subtle symptoms, which complicates effective treatment(20). Ovarian cancer has a 5-year survival rate of roughly 47%, mainly as a result of recurrence and chemotherapy resistance(21). Despite the indisputable fact that novel therapies, such as targeted therapy and immunotherapy, have been continuously emerging in recent years, current treatments for ovarian cancer are still limited(22). Hence, finding particular diagnostic indicators and viable treatment approaches for ovarian cancer is crucial. The m6A modification is the most frequent internal change in eukaryotic mRNA, significantly influencing tumorigenesis and various other biological activities (9). In this study, although mutations in m6A regulatory genes are rare in ovarian cancer, these genes are abnormally expressed. The expression of 23 m6A-gene regulators in ovarian cancer was further shown to differ generally and positively correlated. In addition, based on the collected gene sets of 12 programmed cell deaths, we speculate that the m6A regulatory genes and autophagy were strongly linked in ovarian cancer based on GSVA analysis. Univariate cox regression analysis of autophagy-related genes and m6A regulators showed that there were 36 genes linked to the prognosis of ovarian cancer. LASSO regression analysis was carried out using these 36 genes, and the results demonstrated that the prognostic characteristics of FOXK1, FOXO1, HTR2B, LAMP3, LAMTOR5, LEPR, NPC1, PLEKHF1, PLK2, and RNF5 in ovarian cancer patients were predictive of OS in ovarian cancer patients. Next, depending on the expression levels of these 10 important genes, a risk score (high risk vs. low risk) is established. This model effectively predicts survival outcomes of patients with ovarian cancer, revealing a significant survival difference between the high-risk and low-risk groups.

**Figure 5.**
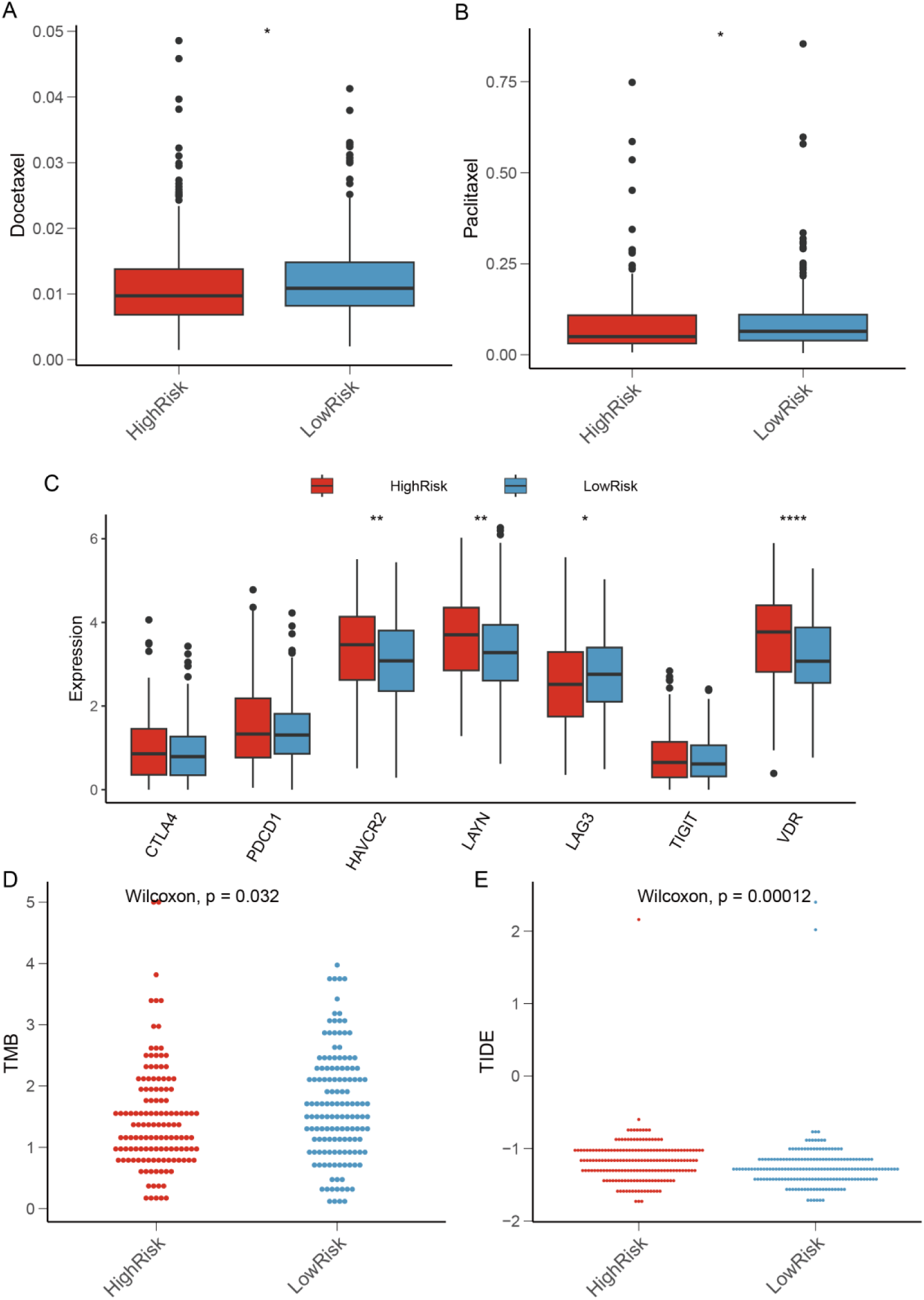
(A-C) Drug sensitivity to Docetaxel and Paclitaxel in high- and low-risk groups. The autophagy high/low risk score patients’ TMB and TDE values in (D) and (E), respectively.

**Figure 6.**
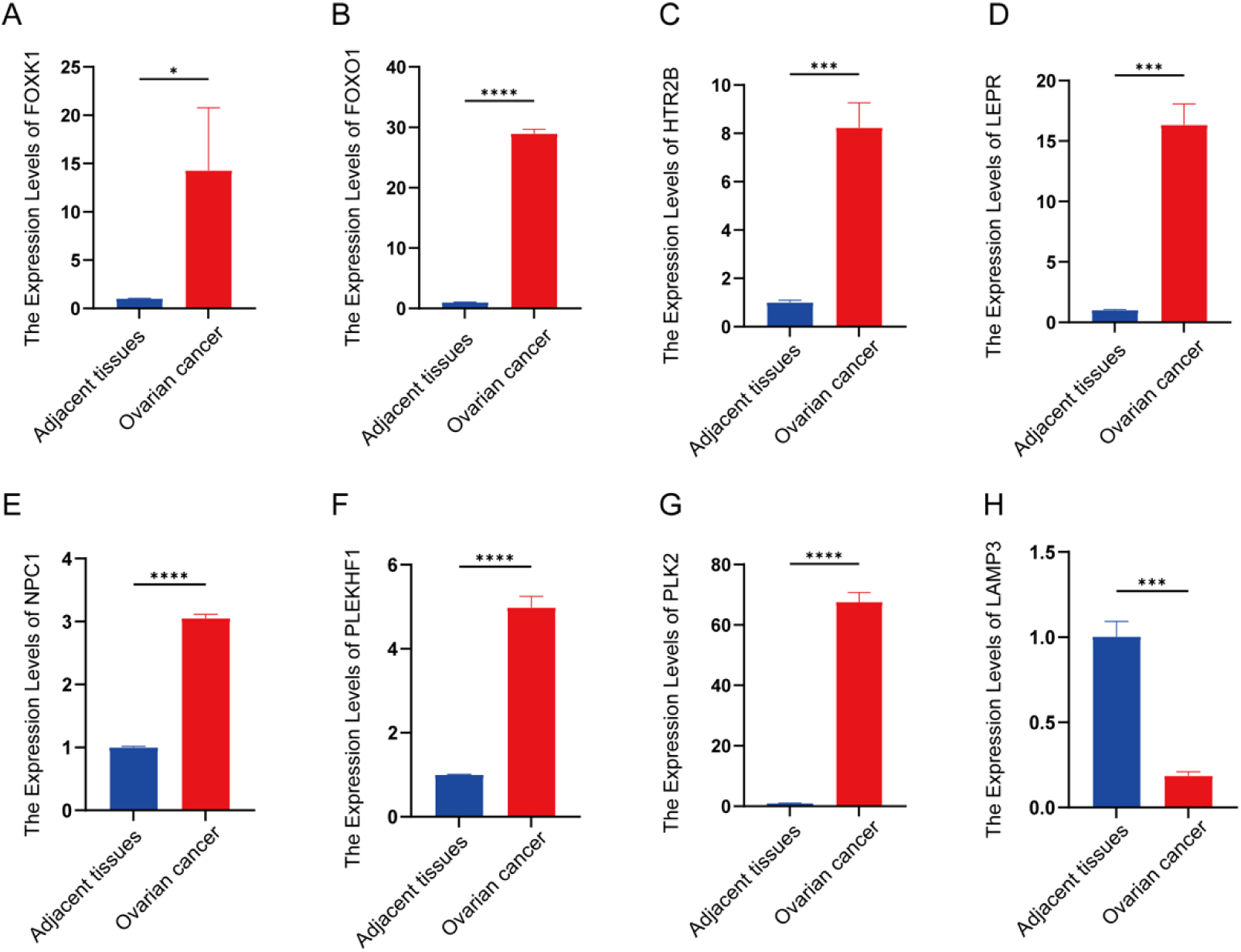
RT-qPCR was used to evaluate the expression of m6A-related genes, such as FOXK1 (A), FOXO1 (B), HTR2B (C), LEPR (D), NPC1 (E), PLEKHF1 (F), PLK2 (G), and LAMP3 (H), in ovarian cancer (n = 4) and neighboring normal tissues (n = 4). The paired sample t test was used to assess the data. The data is displayed in the form of mean ± standard deviation (SD). *p< 0.05, **p< 0.01, ***p< 0.001, ****p< 0.0001, ns, not significant.

**Figure 7.**
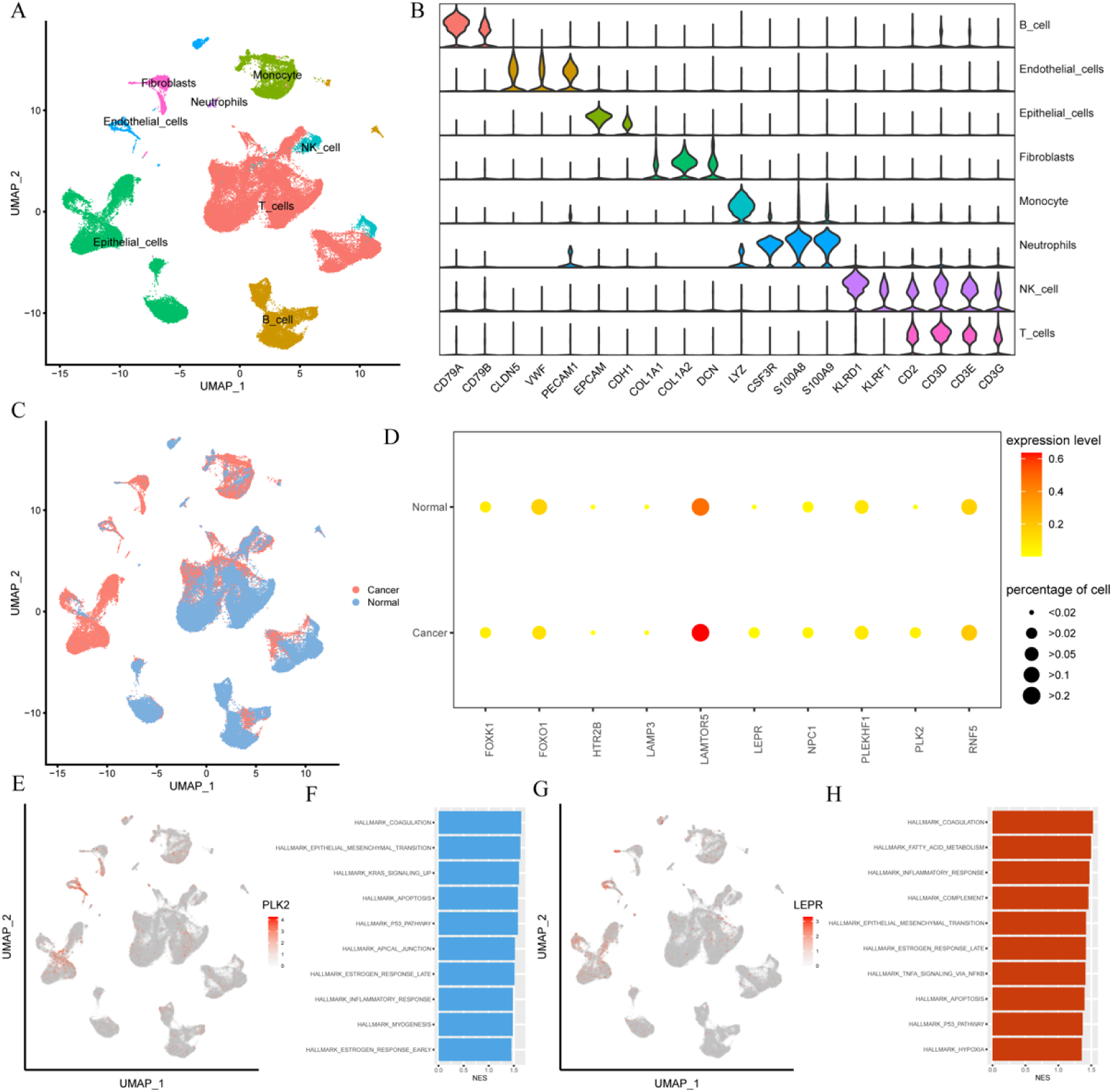
Single-cell RNA sequence analysis shows the expression of key feature across different cell types in ovarian cancer samples. (A) Cell types in GSE184362 are depicted in the Uniform Manifold Approximation and Projection (UMAP) plot. (B) Violin plots displaying the expression level of markers across various cell types. (C) UMAP plot showing the cells from different tissues. (D) The proportion of cells expressing 10 key feature genes and their expression levels in ovarian cancer and adjacent non-tumor samples. (E-H) The UMAP plot and Functional enrichment of PLK2 and LEPR.

It has been found that FOXK1 expression is upregulated in ovarian cancer, and FOXK1 significantly improves cell proliferation and metastasis by regulating the expression of p21 protein and promoting cell cycle(23). FOXO1 is a crucial transcription factor whose knockdown reduces the proliferation of ovarian cancer cells both in vitro and in vivo. Additionally, structural maintenance of chromosome 4 (SMC4), a gene transcriptionally regulated by FOXO1, mediates the exacerbation of ovarian cancer malignant phenotype by FOXO1 in vivo. Notably, FOXO1 also promotes SMC4 mRNA stability through METTL14-mediated m6A modification. These findings offer novel insights into the pathogenesis of ovarian cancer(24). Recent research has highlighted the significant role of 5-Hydroxytryptamine (5-HT) signaling in tumor growth. Specifically, human gastric adenocarcinoma tissues have greater concentrations of the receptor HTR2B than non-tumor tissues do, and higher levels of HTR2B are associated with shorter patient survival times. Reducing or blocking HTR2B with selective antagonists has shown to suppress tumor growth(25). Thus, HTR2B expression could serve as a promising prognostic biomarker for gastric cancer patients. Lysosome-associated membrane protein 3 (LAMP3) has profound implications in the advancement of various gynecological cancers, such as those affecting the cervix, breast, and ovaries. Research indicates that LAMP3 can influence the progression and outcome of Uterine corpus endometrial carcinoma (UCEC) by modulating immune infiltration levels and interacting with chemokines, their receptors, immune enhancers, immunosuppressants, and MHC molecules(26, 27). During malignancy, disordered O-glycosylation has a global impact on disease progression. In breast cancer metastases, abnormal O-glycosylation initiation is triggered by LAMTOR5. LAMTOR5 is significantly upregulated in adenocarcinomas and correlates with Tn antigens, the products of O-glycosylation initiation, both in clinical samples from metastatic breast cancer as well as in mouse models of secondary metastasis(28).

Leptin stimulation enhanced the proliferation, survival and migration of ovarian cancer cell lines expressing LEPR, with these effects mediated by the downstream Janus kinase 2/Signal transducer and activator of transcription 3 (JAK2/STAT3) pathway(29). Cancer cells rely on cholesterol, and increased Niemann Pick C1 (NPC1) facilitates the uptake of extracellular cholesterol, which is essential for the invasion of ErbB2-expressing breast cancer spheroids and ovarian cancer organoids (30). This suggests that NPC1 plays a regulatory role in the process. Polo-like kinase Plk2 is a key factor in determining chemosensitivity, highlighting its potential as a theranostic marker for managing epithelial ovarian cancer (EOC)(31). Additionally, inhibiting RNF5 enhances adhesion and reduces migration in HER2-negative breast cancer cells, while silencing RNF5 limits the growth of xenograft tumors from ER-positive, HER2-negative breast cancer cells, which exhibit increased Ephrin receptor A2 (EphA2) expression and altered phosphorylation(32).

The predictive capability of these ten key genes together is significantly higher than that of any individual factor. The ROC curve showed robust predictive accuracy at 5 years, and the risk score model combined with clinical characteristics (age, stage) could offer enhanced prognostic predictions for ovarian cancer patients. For clinical applications, this raises the prognostic feature’s usefulness.

## 5. Conclusions

This study analyzed the association between m6A and autophagy. In conclusion, our study successfully identified 10 key genes associated with both autophagy and m6A in ovarian cancer patients, which are linked to survival outcomes in ovarian cancer patients. This study emphasizes the significance of RNA modifications in ovarian cancer development and offers potential biomarkers for treatment selection and prognosis evaluation in these patients.

## Declarations

### Consent for publication

**Informed consent was obtained from all subjects involved in the study.**

### Availability of data and materials

The datasets used and/or analysed during the current study are available from the corresponding author on reasonable request.

### Supplementary Materials

The following supporting information can be downloaded at: Figure S1: The overall mutation rate of 23 m6A regulators in ovarian cancer; Figure S2: Building the m6A prognostic model for ovarian cancer. (A) Using univariable Cox regression and a forest plot, m6A regulatory genes associated with ovarian cancer prognosis were found. (B) Cross-validation was used in the LASSO Cox regression to fine-tune the parameter selection. (C) Analysis of m6A regulatory genes through LASSO Cox regression; Figure S3: The immumohistochemical staining of FTO, FOXO1, HTR2B and PLK2 from Human Protein Atlas database.

### Author Contributions

Conceptualization, L.G. and N.H.; methodology, S.S.; software, Y.C.; validation, Y.C., X.Y. and S.S.; writing—original draft preparation, X.X.; writing—review and editing, Y.C.; project administration, C.W.; funding acquisition, N.H. and L.G. All authors have read and agreed to the published version of the manuscript.

### Institutional Review Board Statement

The study was conducted in accordance with the Declaration of Helsinki, and approved by the Institutional Review Board of Qingdao University (QDU-HEC-2024261).

### Informed Consent Statement

Written informed consent has been obtained from the patients to publish this paper.

### Data Availability Statement

The datasets presented in this study can be found in online repositories. These datasets were downloaded from TCGA database, GSE138866, GSE119168 and GSE184362.

## Acknowledgments

We thank the members of the research team for their support and guidance.

